# Computer-delivered Cognitive Training and Transcranial Direct Current Stimulation in Patients with HIV-associated Neurocognitive Disorder: A Randomized Trial

**DOI:** 10.1101/2021.08.22.21262416

**Authors:** Raymond L. Ownby, Jae Kim

**Affiliations:** Department of Psychiatry and Behavioral Medicine, Nova Southeastern University, Fort Lauderdale, Florida, USA

**Author notes:** **Correspondence:** Dr. Ownby.

**Keywords:** transcranial direct current stimulation, computer-delivered cognitive training, human immunodeficiency virus, cognition, HIV-associated neurocognitive disorder, mild neurocognitive disorder

## Abstract

**Objective:** HIV infection is associated with impaired cognition, and as individuals grow older, they may also experience age-related changes in mental abilities. Previous studies have shown that computer-based cognitive training (CCT) and transcranial direct current stimulation (tDCS) may be useful in improving cognition in older persons. This study evaluated the acceptability of CCT and tDCS to older adults with HIV-associated neurocognitive disorder, and assessed their impact on reaction time, attention, and psychomotor speed.

**Methods:** In a single-blind randomized study, 46 individuals with HIV-associated mild neurocognitive disorder completed neuropsychological assessments and six 20-minute training sessions to which they had been randomly assigned to one of the following conditions: (1) CCT with active tDCS; (2) CCT with sham tDCS, or (3) watching educational videos with sham tDCS. Immediately after training and again one month later, participants completed follow-up assessments. Outcomes were evaluated via repeated measures mixed effects models.

**Results:** Participant ratings of the intervention were positive. Effects on reaction time were not significant, but measures of attention and psychomotor speed suggested positive effects of the intervention.

**Conclusion:** Both CCT and tDCS were highly acceptable to older persons with HIV infection. CCT and tDCS may improve cognitive in affected individuals.

Registered at ClinicalTrials.gov (NCT03440840).

## 1 Introduction

While there has been significant progress in the treatment of HIV infection using multiple antiretroviral medications, HIV-association neurocognitive disorders (HANDs) continue to be seen in affected individuals, even when their viral loads are nondetectable (Heaton et al., 2010). HANDs are clinically significant because of their impact on patients’ everyday functioning [1-3], medication adherence [1, 4, 5], and quality of life [6-9]. In older persons, HANDs may have an additive or even synergistic effect in older persons, combining the influences of chronic HIV infection and cognitive aging [10, 11].

Few treatments are available for HAND. Stimulants can improve cognition in HAND but may be abused by vulnerable individuals and have undesirable adverse effects [12]. Other medications treatments have been studied, but none has demonstrated clear efficacy [13-18]. Other researchers have suggested that computer-delivered cognitive training (CCT) may be useful in HAND [19-21], but specialized CCT software is not always readily available or affordable. Further, many programs created for CCT are not inherently interesting, reducing users’ motivations for continued use after completing a study for which they were compensated. Another approach may be to use computer gaming software that is already available as a CCT intervention [22, 23]. CCT software developers have sought to increase the inherent interest in their programs through gamification [24] to enhance their inherent interest, but many computer games are on the market now and often available at little or no cost. In addition, existing games depend on sustained use by players for their commercial success. Games such as these are interesting to players and include elements that engage them. First-person shooters (in which players use weapons to shoot at fictional enemies) can affect sustained attention and reaction time [24], however, some players may object to this type of game’s violent content [24, 25].

Another established genre are games that provide players the simulated experience of car racing. These games require attention and psychomotor speed while using content that may be less objectionable. Use of one car racing game, created for a research study, was associated with better mental functioning in older persons [26]. Other researchers have commented on the possible usefulness of commercial computer games in addressing mental functioning in persons 50 years of age and older [23, 27-30]. Car racing games can engage and hold players’ interest, potentially allowing them to continue cognitive training over extended periods. Researchers have shown that an off-the-shelf game that demanded mental speed resulted in longer use by older individuals when compared to a typical CCT program [29, 30]. Game play has been related to long-term mental training results, with effects evident in other cognitive domains besides those specifically trained [31]. Gaming has been shown, for example, to have a positive impact on the ability to regulate and direct mental processes [26]. Games may thus be effective for training and can engage users in a sustained fashion.

Transcranial direct current stimulation (tDCS) in combination with CCT has been shown to improve cognitive functioning [32-38]. tDCS is implemented by applying moistened sponge electrodes to a person’s scalp and passing through a very small direct current (1-2 mA). tDCS research has shown that it can have a positive impact on various mental abilities, including verbal problem solving [39], working memory [40-42], and learning [43, 44].

How tDCS affects mental function is not definitively established, however, it has been shown to stimulate brain-derived neurotrophic growth factor (BDNF) in the motor cortex [45]. This may be especially relevant in treating persons with HAND as BDNF is affected in HIV infection [46, 47], and implicated in cognitive decline in older persons [48]. Increases in BDNF might be expected to exert a positive effect on mental functioning in persons with HAND.

We previously completed a pilot study of game-based CCT comparing its combination with active and sham tDCS in persons 50 years and older with HAND [49]. Results suggested that the intervention was acceptable to participants and that it may have had positive effects on their attention and working memory. In the follow-up study reported here, we further explore the acceptability and efficacy of a game-based CCT intervention combined with tDCS in older persons with HIV infection. We hypothesized that CCT with tDCS would be acceptable to persons 50 years of age and older with HAND. We also hypothesized that CCT would be associated with improved reaction time, psychomotor speed, and attention and that the combination of active tDCS with CCT would be superior to CCT alone.

## 2 Methods

### 2.1 Participants

Participants were individuals 50 years of age and older with HIV infection. Diagnosis of HAND was established through review of recent laboratory results, clinical evaluation, and neuropsychological testing. All participants stated they subjectively experienced cognitive difficulties and, after assessment, were found to have impairment of mental functioning in two or more cognitive domains while not having dementia, thus meeting Frascati criteria for mild neurocognitive disorder [15]. Potential participants were excluded if they had characteristics that might have increased risk to them from tDCS, such as seizures or bipolar disorder [50, 51]. Use of many psychotropic medications was also an exclusion criterion, as the pharmacologic activity of many of these drugs can affect tDCS [52, 53]. Medications that were exclusions included those affecting serotonin, such as many antidepressants, dopamine, such as stimulants and antipsychotics, and gamma-amino butyric acid, such as benzodiazepines. Left-handed participants were excluded as our intent was to stimulate the dominant dorsolateral prefrontal cortex.

### 2.2 Procedures

#### 2.2.1 Recruitment and determination of eligibility

Participants were first recruited from individuals who had been in a previous study. We also recruited from local service providers for persons with HIV. A number of participants referred friends or acquaintances. We distributed flyers in several areas of Broward County, Florida, known to have a high prevalence of HIV infection, as well as advertising in a local newspaper and creating a Facebook page.

Interested individuals were contacted for a telephone interview to establish that they had complaints of cognitive difficulties, using questions published by the European AIDS Clinical Society [54]. In this interview, we inquired about use of medications that might lead to exclusion, and whether the person was willing to be in a study of CCT and tDCS. All were being treated for HIV infection and had been on their current medication regimen for at least the past month. Individuals who, from this telephone interview, appeared likely to be eligible were asked to come to our offices for individual assessment.

At this assessment, potential participants completed a series of cognitive assessments (marked with an asterisk in Table 1). The battery was selected to allow evaluation of areas often affected in HANDs [55]. Attention and working memory were evaluated with the Digit Span subtests of the Wechsler Adult Intelligence Scale, 4th edition, or WAIS-IV [56]. Psychomotor speed was evaluated with the Coding subtest of the WAIS-IV and Grooved Pegboard Test [57]. Executive function was measured with the Trail Making Test, Part B [58]. Verbal fluency was assessed with the Verbal Fluency test of the Delis-Kaplan Executive Function System [59]. Verbal learning and memory were assessed with the Hopkins Verbal Learning Test—Revised or HVLT-R [60], and visual learning and memory with the Brief Visuospatial Memory Test, or BVMT-R [61].

**Table 1.** Cognitive and Functional Measures Used

| Domain | Measure |
| --- | --- |
| Reaction Time | *California Computerized Assessment Package (CalCap)* [66] |
| Attention | * *Wechsler Adult Intelligence Scale, 4<sup>th</sup> Ed.*, Digit Span subtest [99] |
|  | *Wechsler Memory Scale, 4<sup>th</sup> ed.*, Symbol Span subtest [101] |
| Psychomotor Speed | * *Wechsler Adult Intelligence Scale, 4<sup>th</sup> ed.*, Coding subtest [100] |
|  | *Trail Making Test, Part A [78] |
|  | *Grooved Pegboard [51] |
| Premorbid function | Wechsler Test of Adult Reading [98] |
| Executive function | *Trail Making Test, Part B [78] |
|  | *Verbal and Design Fluency from D-KEFS [36] |
|  | Stroop Color Word Test [42] |
|  | Iowa Gambling Task [12] |
| Learning and Memory | *Hopkins Verbal Learning Test – Revised [15] |
|  | *Brief Visuospatial Memory Test – Revised [16] |
| Functional Status | Medication Management Test – Revised [2] |
|  | University of California San Diego Performance-based Skills Assessment [74, 70] |
| *Note: Measures used to establish eligibility are marked with an asterisk. |  |

Cognitive impairment for the purpose of establishing the diagnosis of mild neurocognitive disorder was defined as a score in at least two ability areas that was below population norms by at least one standard deviation. Participants were treated for HIV infection that included ongoing laboratory measures of treatment effects (HIV-1 viral load and CD4 cell counts). Individuals in the study brought recent laboratory results, allowing us to verify their HIV status and know their current treatment and immune status. All medications were also brought to this visit to allow verification of current medication use. Persons who met entry criteria then completed the additional assessments as described in the next section.

#### 2.2.2 Acceptability

We used several strategies to evaluate the feasibility and acceptability of the CCT with tDCS intervention to participants. We used a questionnaire based on the Technology Acceptance Model, or TAM [62, 63], the dimensions of which have received substantial support for use with digital health technologies [64]. The model specifies that users’ perceptions of an application’s ease of use and usefulness are related to their future intention to use the application. We hypothesized that if the intervention were viewed favorably by participants, their average rating on the Usefulness and Ease of Use scales of this questionnaire would be significantly different from the midpoint of the scale in a positive direction.

Another scale was developed based on a model balancing risks and benefits of a treatment was used to develop a questionnaire assessing users’ perceptions of the balance between an intervention’s risks and benefits [65, 66]. Participants were asked, for example, if they experienced benefits from the intervention and adverse effects from it. They were then asked to provide an overall judgment as to whether the benefits of the intervention outweighed its adverse effects.

#### 2.2.3 Cognitive measures

In order to evaluate possible cognitive effects of the intervention more comprehensively, participants additional assessments after determination of their eligibility. Use of these measures allowed tests of the study’s hypothesis that participants receiving the active interventions would display better performance than control participants in reaction time, attention, and psychomotor speed.

In order to evaluate intervention effects on participants’ reaction time, they completed the California Computerized Assessment Package [67] (Miller, 2013). To further evaluate the effects of the intervention on executive functions, participants also completed the Stroop Color Word Test [68], the Iowa Gambling Task [69], and the Design Fluency subtest of the Delis-Kaplan Executive Function System [59](Delis et al., 2001). Finally, to assess whether the intervention had an impact on everyday functional performance, participants completed the Medication Management Test—Revised, a measure of the person’s ability to understand and carry out medication-related tasks [70] and the University of San Diego Scales of Observed Performance [71], assessing their ability to perform everyday tasks such as making a medical appointment and paying a bill. Finally, in order to provide an estimate of participants’ premorbid level of functioning, they completed the Wechsler Test of Adult Reading [72].

#### 2.2.4 Other self-report measures

The assessment battery also included the Patient’s Assessment of Own Functioning or PAOF [73]. This measure asks the individual to self-report their experience of mental problems in several domains such as language, perception, and memory. It has been used in other studies of HAND [74]. We also used the Center for Epidemiological Studies Depression scale or CESD [75] to assess participants’ symptoms of depression. All self-report assessments were completed using computer software that read questions aloud and enabled participants to record their responses by tapping on the computer screen.

#### 2.2.5 Compensation

After these initial assessments were done, individuals in the study were asked to return to begin the intervention. Participants received compensation for their involvement, US $80 for the baseline and follow-up sessions, and $40 for each intervention visit.

#### 2.2.6 Computer-based cognitive training

At the first training visit (after completion of baseline assessments), participants were assigned to intervention group using a computer-created randomization scheme. The scheme was generated via random numbers in a predetermined block (n = 3) randomization scheme. Participants were enrolled by the study coordinator (who was blind to treatment assignment) and assigned by the unblinded principal investigator who also conducted all training sessions.

First, procedures regarding the administration of tDCS and the use of the game controller (an Xbox game controller with USB interface to a Windows computer). The participant sat in front of and to the right of the researcher; the participant could not see the direct current device for tDCS or the researcher taking notes during training. For all participants, the anode electrode was located over the left dorsolateral prefrontal cortex (10-20 system F3) and the cathode over the right supraorbital area (FP2) [76]. Soterix EASYPads, doubled sponges with dimensions of 5 cm x 5 cm (Soterix Medical: New York) were used as electrodes. Approximately 7 cc of sterile saline was used to moisten them. They were positioned by the researcher and then fixed in place with a head band. Current for the tDCS intervention was supplied with an iontophoresis device (ActivaDose II; Gilroy, CA: Activatek). Flat rubberized carbon electrodes were inserted into the moistened sponges. Impedances were assessed before each session and kept below 20K ohms prior to stimulation.

We told people in the study that they might experience minor discomfort at the beginning of the session and that the experience might persist or fade away during the intervention [50]. We then asked the persons in the study to pay attention to the computer while the game was set up and the tDCS intervention was begun. Individuals in the active tDCS group received a current of 1.5 mA, ramping up over 30 s and continuing for 20 minutes. Individuals in the sham tDCS group and the control group received the ramping up current for 30 s which was then ramped down over 30 s.

The cognitive training intervention in this study was a commercially-available car racing game *GT Racing 2* (Gameloft SE: Paris, France). This game was chosen because it was easy for most persons and was positively reviewed by a large number of users. We inferred from these characteristics that the participants in the study would find the content tolerable and might even enjoy playing the game. The game includes a several different race courses and types of races to enhance player interest. Game play required that each participant complete each course before moving on to the next. Everyone in the study randomized to CCT was able to complete at least the first four courses during six training sessions.

We encouraged participants to complete each gaming session at their desired pace, although the game imposed some restrictions on their progress. Players had to finish each race in one of the top three places, or navigate a course within a predetermined time, before moving on to the next course. We required as well that participants do each course a minimum of five times. Persons in the study completed six training sessions over a two-week period. After each intervention session, participants were asked to provide ratings of their thinking, their mood, and how much discomfort they had experienced during the intervention.

Assessments and intervention sessions were completed within three weeks from the baseline evaluation. After participants finished the sixth intervention session, they returned to complete cognitive and self-report assessment. Cognitive evaluations were completed by staff who did not know the participant’s intervention group assignment. About 30 days after completing the intervention sessions and follow-up evaluations, persons in the study were asked to return to again complete assessments. All data were collected in the General Clinical Research Center at the Center for Collaborative Research on the campus of Nova Southeastern University beginning in January 2018 and ending in November 2019. The study was concluded at the end of the period of funding support.

### 2.3 Human subjects approval and trial registration

Study procedures were approved by the Institutional Review Board of Nova Southeastern University (protocol number 2017-410). This study was registered on ClinicalTrials.gov (NCT03440840).

### 2.4 Data Analyses

Planned sample size was determined prior to beginning the study using the mixed effects model simulation routine in PASS 16 [77]. The power analysis showed that a final sample size of 90 (30 per treatment group) would have a power of 0.88 to detect interactions of group membership with time (number of evaluations) with a small effect size [78, 79].

Data analyses were completed in several steps. Preliminary analyses of data and descriptive statistics were obtained using SPSS version 26 (Armonk NY: IBM). Chi-square and one-way ANOVA tests evaluating the relations of participant ratings of the acceptability and feasibility of the intervention to group assignment were also completed in SPSS. Analyses of treatment effects were completed using R version 4.0.2 [80] package lme4 [81] for mixed effects models. Significance of model effects (interaction of treatment group assignment with time) was assessed using the likelihood ratio test [82]. We evaluated outcomes both through tests of statistical significance as well as approximations of effect size from χ2 values from likelihood ratio tests and t values obtained in tests of between-group differences obtained using emmeans [83]. Effect sizes were converted to the more familiar d statistic using the package esc [84]. As a post-hoc assessment not included in the original study protocol the probability of finding the observed number of treatment effects in the hypothesized direction was evaluated using the exact binomial test in the package stats.

## 3 Results

The study’s CONSORT diagram [85] is presented in Figure 1. Demographic, educational and baseline scores for cognitive and functional measures for participants by treatment group are presented in Table 2. We screened 155 potential participants, and included 46 in the study. Reasons for excluding potential participants are listed in Figure 1. The most frequent reasons for exclusion were use of psychotropic medications, and a personal or family history of bipolar disorder. We thus were not able to include the full number of participants in the study as originally envisioned (planned N = 30 per group).

**Figure 1.**
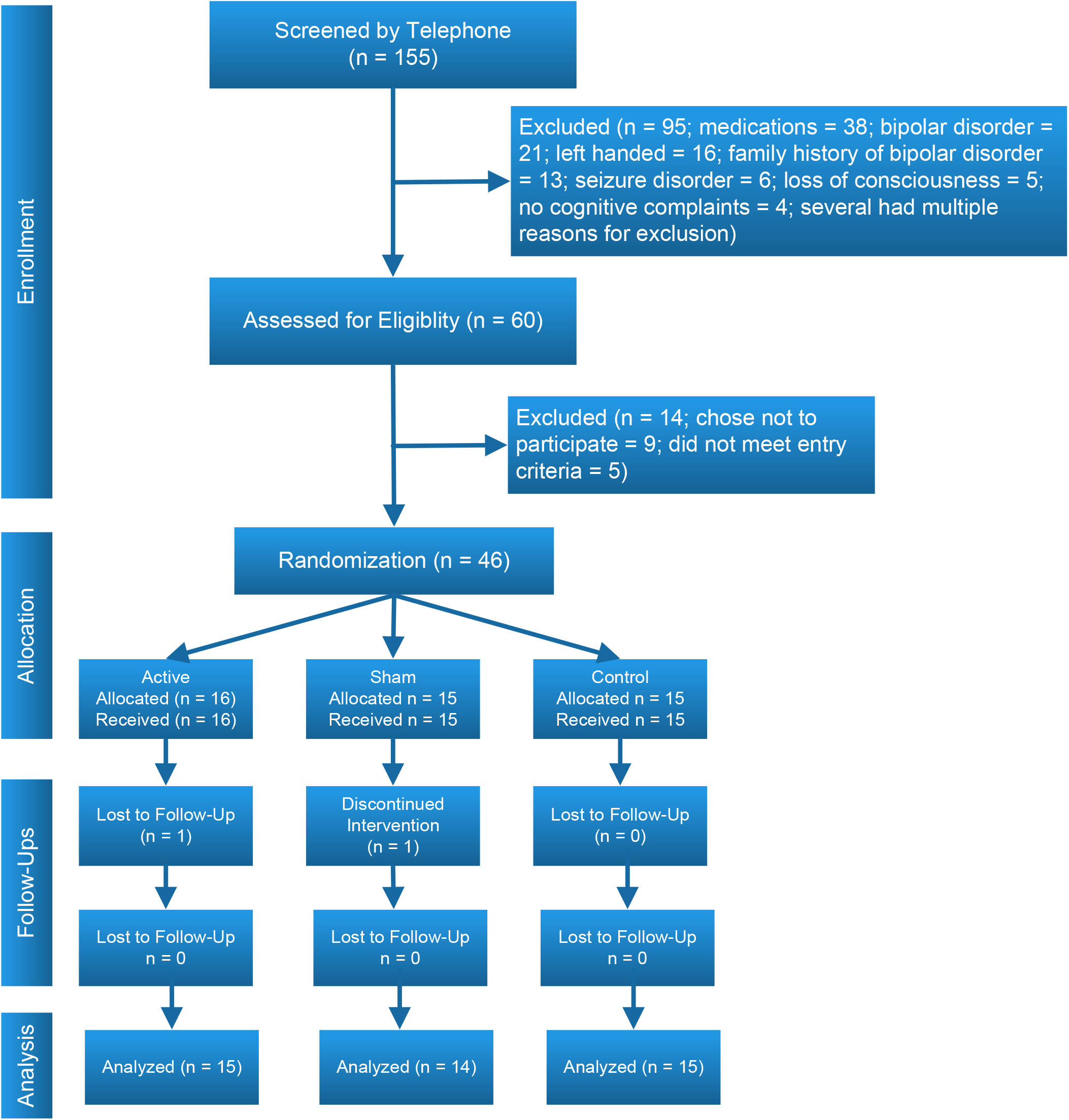
CONSORT diagram

**Table 2.**
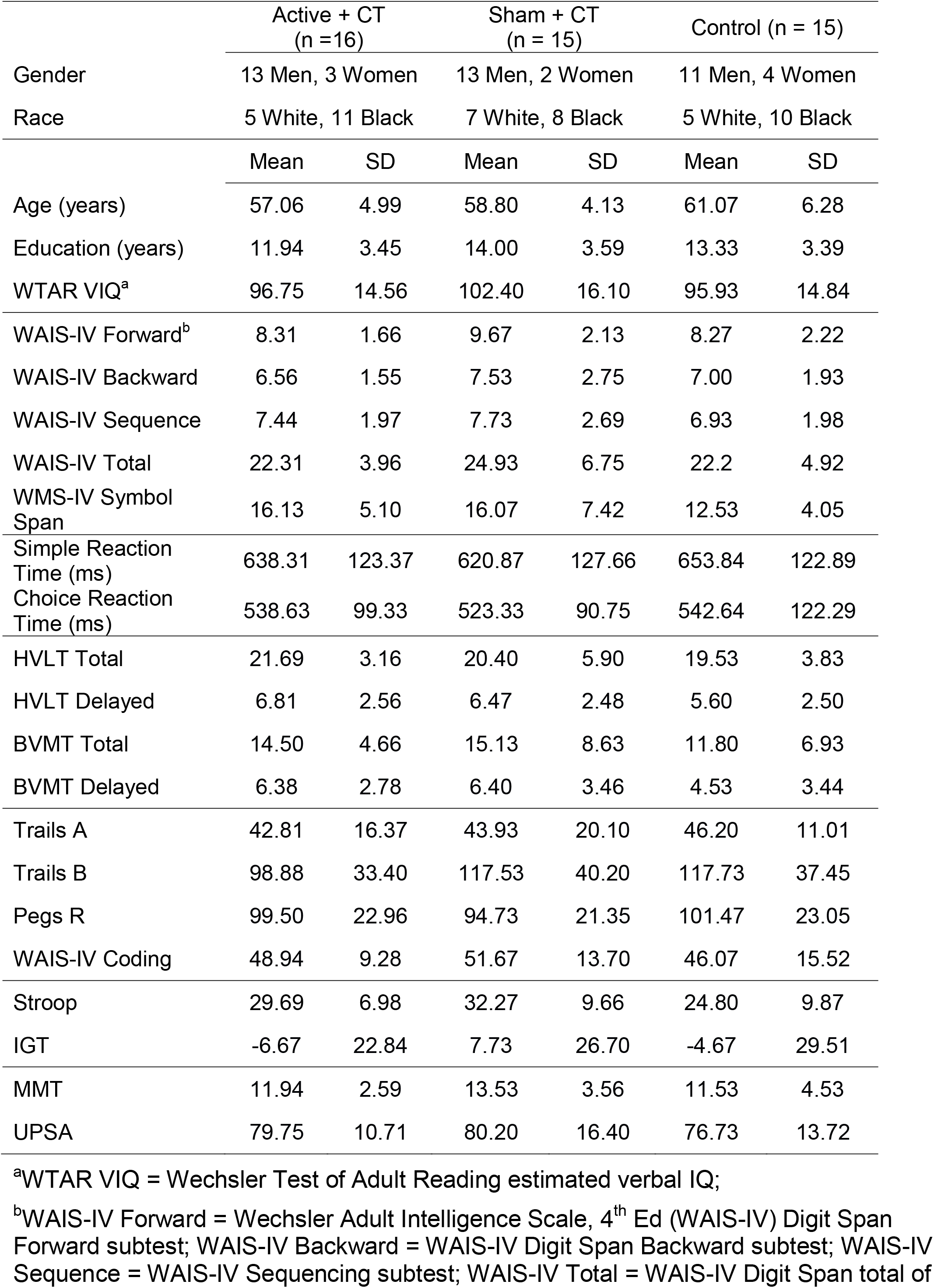
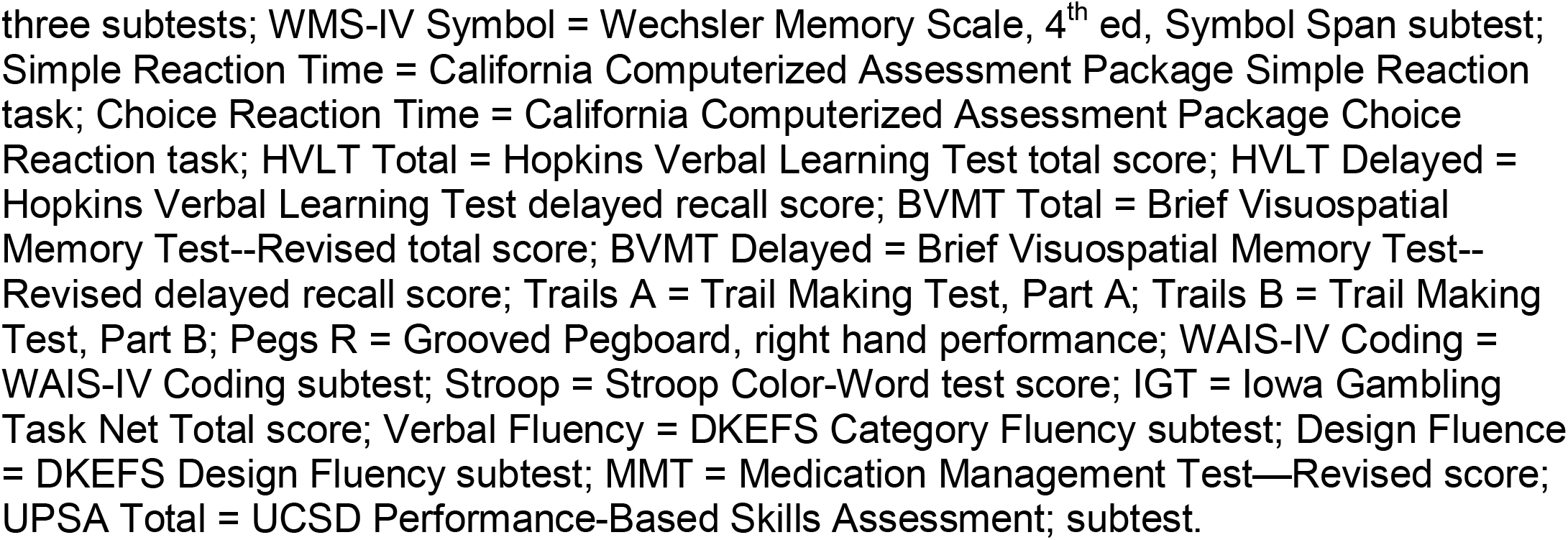
Baseline characteristics of treatment groups

|  | Active + CT<br>(n =16) |  | Sham + CT<br>(n = 15) |  | Control (n = 15) |  |
| --- | --- | --- | --- | --- | --- | --- |
| Gender | 13 Men, 3 Women |  | 13 Men, 2 Women |  | 11 Men, 4 Women |  |
| Race | 5 White, 11 Black |  | 7 White, 8 Black |  | 5 White, 10 Black |  |
|  | Mean | SD | Mean | SD | Mean | SD |
| Age (years) | 57.06 | 4.99 | 58.80 | 4.13 | 61.07 | 6.28 |
| Education (years) | 11.94 | 3.45 | 14.00 | 3.59 | 13.33 | 3.39 |
| WTAR VIQ<sup>a</sup> | 96.75 | 14.56 | 102.40 | 16.10 | 95.93 | 14.84 |
| WAIS-IV Forward<sup>b</sup> | 8.31 | 1.66 | 9.67 | 2.13 | 8.27 | 2.22 |
| WAIS-IV Backward | 6.56 | 1.55 | 7.53 | 2.75 | 7.00 | 1.93 |
| WAIS-IV Sequence | 7.44 | 1.97 | 7.73 | 2.69 | 6.93 | 1.98 |
| WAIS-IV Total | 22.31 | 3.96 | 24.93 | 6.75 | 22.2 | 4.92 |
| WMS-IV Symbol Span | 16.13 | 5.10 | 16.07 | 7.42 | 12.53 | 4.05 |
| Simple Reaction Time (ms) | 638.31 | 123.37 | 620.87 | 127.66 | 653.84 | 122.89 |
| Choice Reaction Time (ms) | 538.63 | 99.33 | 523.33 | 90.75 | 542.64 | 122.29 |
| HVLT Total | 21.69 | 3.16 | 20.40 | 5.90 | 19.53 | 3.83 |
| HVLT Delayed | 6.81 | 2.56 | 6.47 | 2.48 | 5.60 | 2.50 |
| BVMT Total | 14.50 | 4.66 | 15.13 | 8.63 | 11.80 | 6.93 |
| BVMT Delayed | 6.38 | 2.78 | 6.40 | 3.46 | 4.53 | 3.44 |
| Trails A | 42.81 | 16.37 | 43.93 | 20.10 | 46.20 | 11.01 |
| Trails B | 98.88 | 33.40 | 117.53 | 40.20 | 117.73 | 37.45 |
| Pegs R | 99.50 | 22.96 | 94.73 | 21.35 | 101.47 | 23.05 |
| WAIS-IV Coding | 48.94 | 9.28 | 51.67 | 13.70 | 46.07 | 15.52 |
| Stroop | 29.69 | 6.98 | 32.27 | 9.66 | 24.80 | 9.87 |
| IGT | -6.67 | 22.84 | 7.73 | 26.70 | -4.67 | 29.51 |
| MMT | 11.94 | 2.59 | 13.53 | 3.56 | 11.53 | 4.53 |
| UPSA | 79.75 | 10.71 | 80.20 | 16.40 | 76.73 | 13.72 |

<sup>a</sup>WTAR VIQ = Wechsler Test of Adult Reading estimated verbal IQ;
<sup>b</sup>WAIS-IV Forward = Wechsler Adult Intelligence Scale, 4<sup>th</sup> Ed (WAIS-IV) Digit Span Forward subtest; WAIS-IV Backward = WAIS-IV Digit Span Backward subtest; WAIS-IV Sequence = WAIS-IV Sequencing subtest; WAIS-IV Total = WAIS-IV Digit Span total of
three subtests; WMS-IV Symbol = Wechsler Memory Scale, 4<sup>th</sup> ed, Symbol Span subtest; Simple Reaction Time = California Computerized Assessment Package Simple Reaction task; Choice Reaction Time = California Computerized Assessment Package Choice Reaction task; HVLT Total = Hopkins Verbal Learning Test total score; HVLT Delayed = Hopkins Verbal Learning Test delayed recall score; BVMT Total = Brief Visuospatial Memory Test--Revised total score; BVMT Delayed = Brief Visuospatial Memory Test-- Revised delayed recall score; Trails A = Trail Making Test, Part A; Trails B = Trail Making Test, Part B; Pegs R = Grooved Pegboard, right hand performance; WAIS-IV Coding = WAIS-IV Coding subtest; Stroop = Stroop Color-Word test score; IGT = Iowa Gambling Task Net Total score; Verbal Fluency = DKEFS Category Fluency subtest; Design Fluence = DKEFS Design Fluency subtest; MMT = Medication Management Test—Revised score; UPSA Total = UCSD Performance-Based Skills Assessment; subtest.

We explored relations of relevant covariates (age, gender, education, immune status) to mental ability variables with standard measures of association (correlations). As several of these associations were substantial and likely to create confounding relations, they were included as covariates in mixed effects random intercept models. Outcomes assessed were changes in test scores across treatment groups before and after the study intervention.

### 3.1 Acceptability to participants

As hypothesized, participants rated the intervention significantly more positively than the midpoint of the Usefulness subscale of the TAM scale. The mean rating for all participants was 4.33 (SD = 1.27; scale range 0 to 6), and this rating was significantly greater than the neutral midpoint (t [40] = 6.71, p < 0.001). We also found that they rated its ease of use positively with a mean rating of 5.04 (SD = 0.95), ratings that were again significantly greater than the scale midpoint (t [43] = 5.04, p < 0.001). Ratings suggested that overall, they enjoyed the intervention (mean = 4.73; SD = 1.30) and would use it again if given the opportunity (mean = 4.62; SD = 1.25). Participants’ ratings did not vary by treatment group (all *p*s > 0.35).

On the scale which asked participants to assess the intervention’s risks and benefits, participant ratings again suggested a positive evaluation, with a mean rating of 4.53 (SD = 1.20; scale range 0 to 6) on the item “overall the good outweighs the bad” and a rating of 4.70 (SD = 1.01) on the item “overall satisfaction.” Forty-three of 46 participants (93%) indicated they were satisfied with the intervention.

### 3.2 Cognitive and functional outcomes

Results of evaluations of study outcomes assessed as the interactions of intervention group across evaluations are available in Table 3.

**Table 3.**
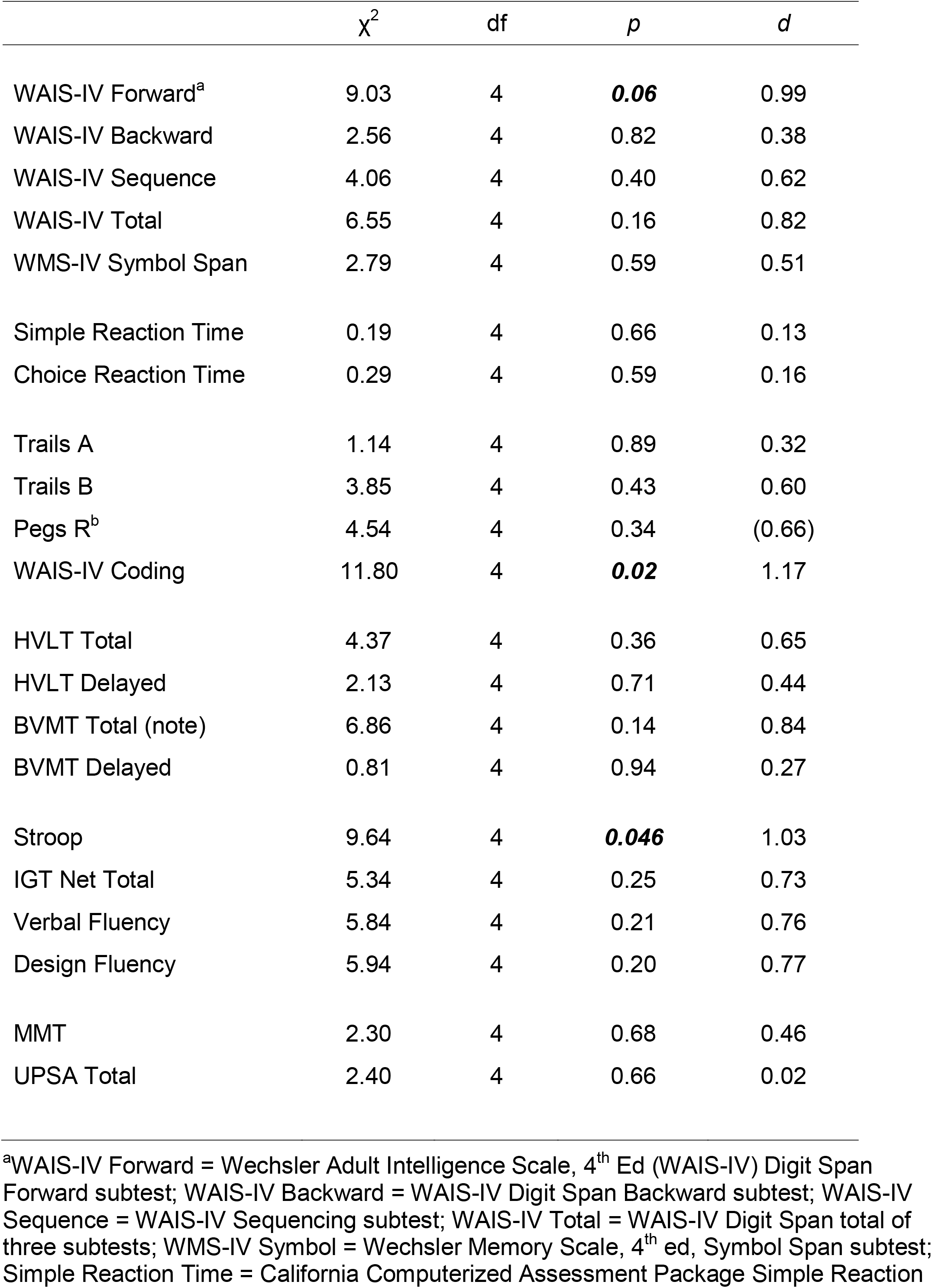
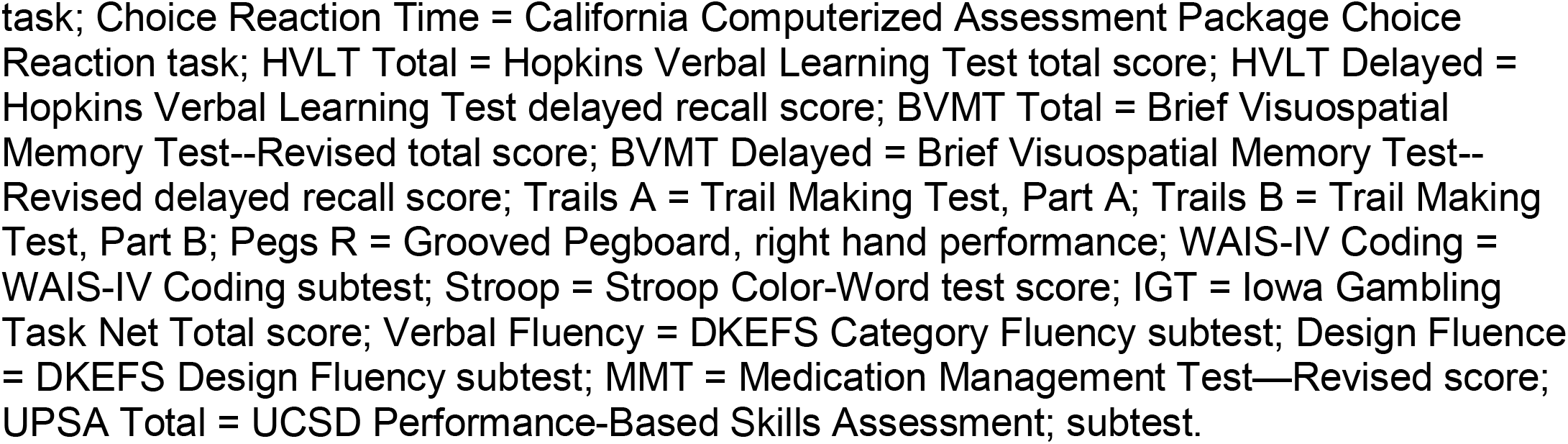
Likelihood ratio test and effect sizes for the interaction of group by time

| | $\chi^2$ | df | *p* | *d* |
| --- | --- | --- | --- | --- |
| WAIS-IV Forward<sup>a</sup> | 9.03 | 4 | **0.06** | 0.99 |
| WAIS-IV Backward | 2.56 | 4 | 0.82 | 0.38 |
| WAIS-IV Sequence | 4.06 | 4 | 0.40 | 0.62 |
| WAIS-IV Total | 6.55 | 4 | 0.16 | 0.82 |
| WMS-IV Symbol Span | 2.79 | 4 | 0.59 | 0.51 |
| Simple Reaction Time | 0.19 | 4 | 0.66 | 0.13 |
| Choice Reaction Time | 0.29 | 4 | 0.59 | 0.16 |
| Trails A | 1.14 | 4 | 0.89 | 0.32 |
| Trails B | 3.85 | 4 | 0.43 | 0.60 |
| Pegs R<sup>b</sup> | 4.54 | 4 | 0.34 | (0.66) |
| WAIS-IV Coding | 11.80 | 4 | **0.02** | 1.17 |
| HVLT Total | 4.37 | 4 | 0.36 | 0.65 |
| HVLT Delayed | 2.13 | 4 | 0.71 | 0.44 |
| BVMT Total (note) | 6.86 | 4 | 0.14 | 0.84 |
| BVMT Delayed | 0.81 | 4 | 0.94 | 0.27 |
| Stroop | 9.64 | 4 | **0.046** | 1.03 |
| IGT Net Total | 5.34 | 4 | 0.25 | 0.73 |
| Verbal Fluency | 5.84 | 4 | 0.21 | 0.76 |
| Design Fluency | 5.94 | 4 | 0.20 | 0.77 |
| MMT | 2.30 | 4 | 0.68 | 0.46 |
| UPSA Total | 2.40 | 4 | 0.66 | 0.02 |

<sup>a</sup>WAIS-IV Forward = Wechsler Adult Intelligence Scale, 4<sup>th</sup> Ed (WAIS-IV) Digit Span Forward subtest; WAIS-IV Backward = WAIS-IV Digit Span Backward subtest; WAIS-IV Sequence = WAIS-IV Sequencing subtest; WAIS-IV Total = WAIS-IV Digit Span total of three subtests; WMS-IV Symbol = Wechsler Memory Scale, 4<sup>th</sup> ed, Symbol Span subtest; Simple Reaction Time = California Computerized Assessment Package Simple Reaction
task; Choice Reaction Time = California Computerized Assessment Package Choice Reaction task; HVLT Total = Hopkins Verbal Learning Test total score; HVLT Delayed = Hopkins Verbal Learning Test delayed recall score; BVMT Total = Brief Visuospatial Memory Test--Revised total score; BVMT Delayed = Brief Visuospatial Memory Test-- Revised delayed recall score; Trails A = Trail Making Test, Part A; Trails B = Trail Making Test, Part B; Pegs R = Grooved Pegboard, right hand performance; WAIS-IV Coding = WAIS-IV Coding subtest; Stroop = Stroop Color-Word test score; IGT = Iowa Gambling Task Net Total score; Verbal Fluency = DKEFS Category Fluency subtest; Design Fluence = DKEFS Design Fluency subtest; MMT = Medication Management Test—Revised score; UPSA Total = UCSD Performance-Based Skills Assessment; subtest.

We found mixed evidence across cognitive domains to support the hypothesis that CCT with and without tDCS might result in improvements in cognitive functioning relative to control. For the Digit Span Forward subtest, the interaction of treatment group by time approached statistical significance and represented a large effect size. The difference between active tDCS and the control group (Figure 2) also approached statistical significance (t [61.3] = 2.19, p = 0.08; d = 0.79). Other subtests hypothesized to assess attention showed similar positive but nonsignificant interactions.

**Figure 2.**
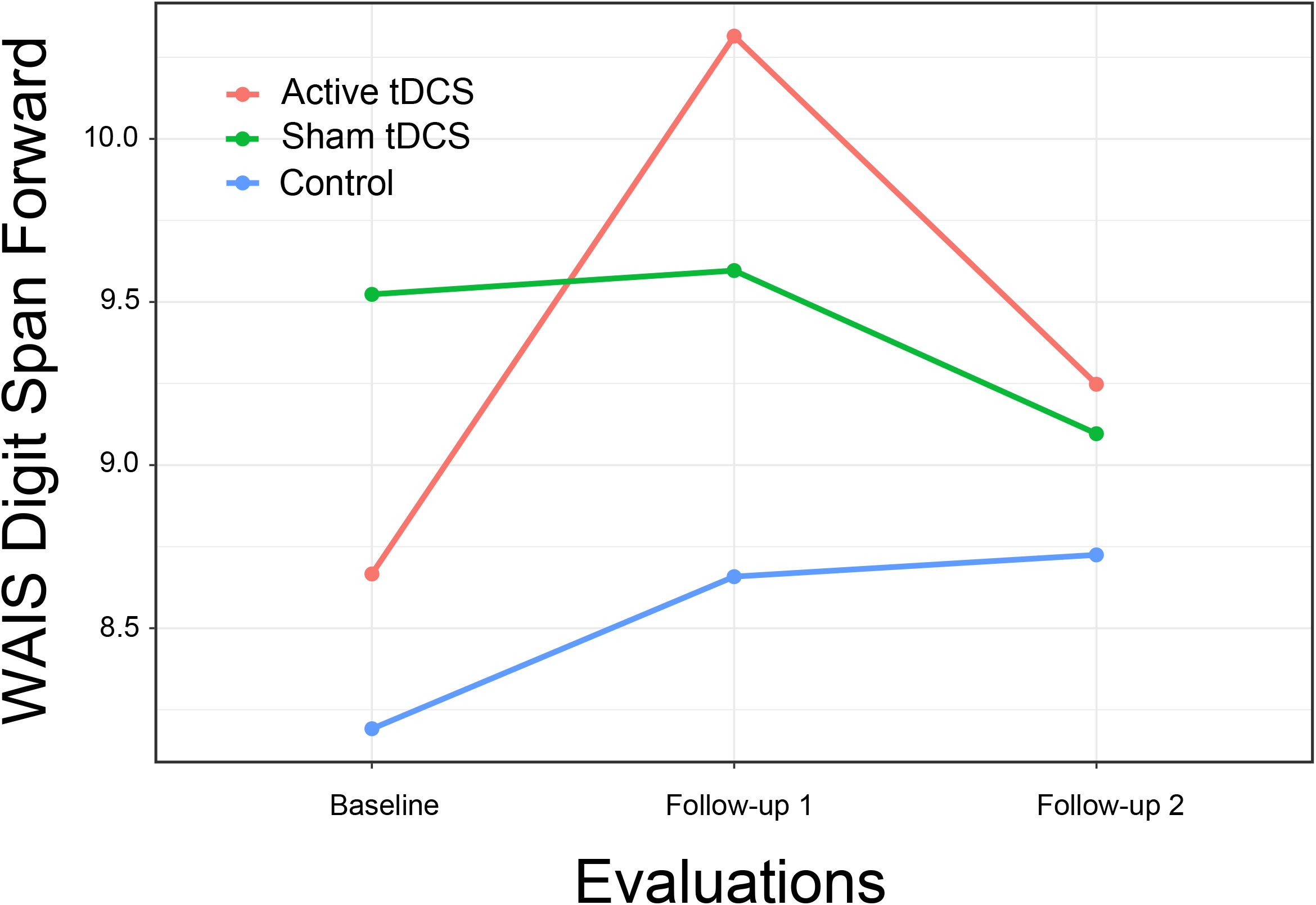
WAIS-IV Digit Span Forward subtest score by Group and Time

Results did not support the hypothesis that persons receiving CCT with or without tDCS would show improved reaction time. Although the observed interactions were in the hypothesized direction, they were not significant and represented at best a very small effect size.

Results again provided limited support for the hypothesis that CCT with tDCS might result in improved psychomotor speed compared to control (Figure 3). Although the overall interaction of group by time was not significant for Trails B, the comparison of the active treatment group to control approached significance (t [67.2] = 2.37, p = 0.053; d = 0.85), a difference that became significant at second follow-up (t [67.2] = 3.03, p = 0.01; d = 1.09).

**Figure 3.**
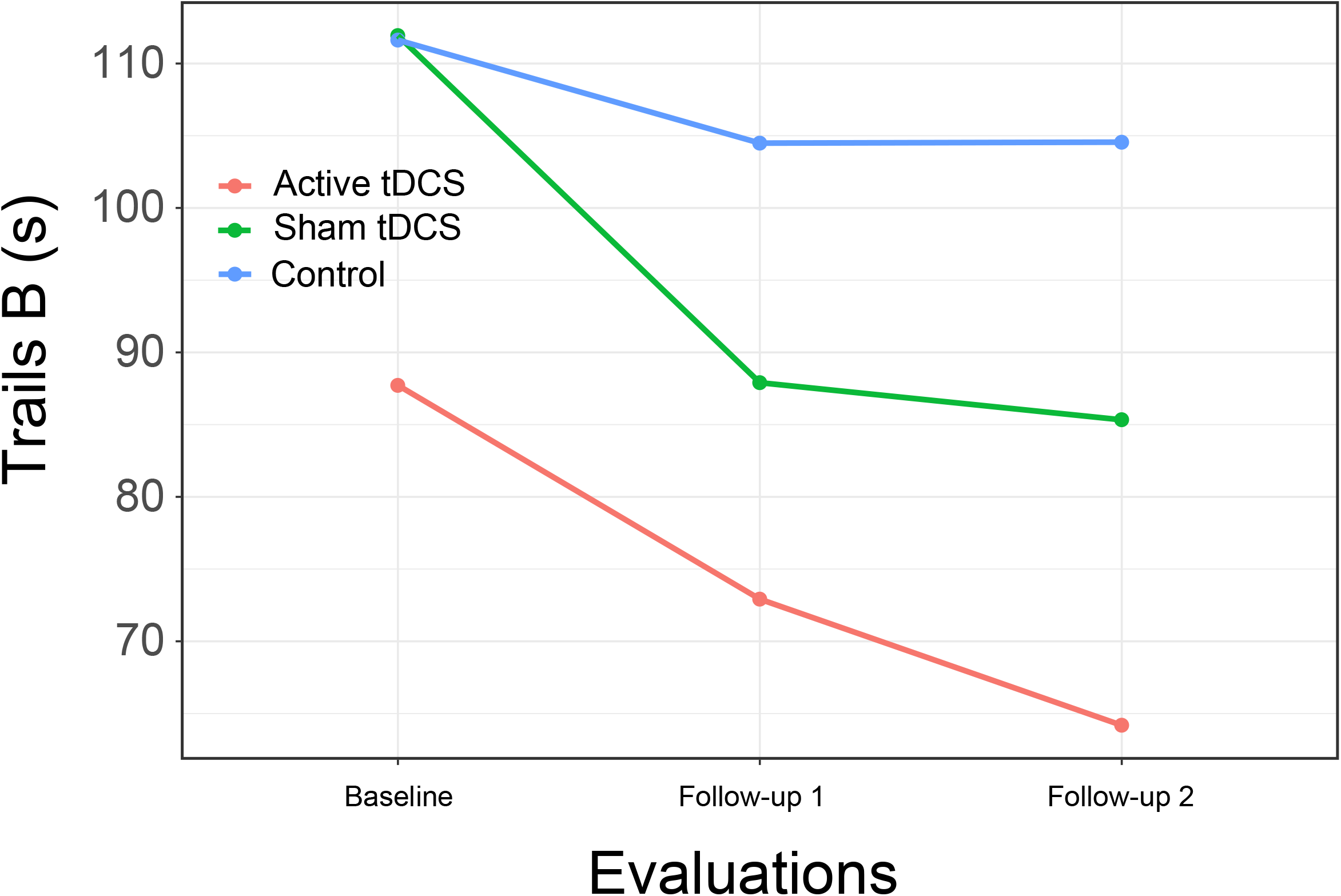
Trail Making Test, Part B by Group and Time Figure 3 footnote: Lower scores indicate better performance

We found support for the hypothesis of improvement in psychomotor speed with a significant interaction of group by time for the WAIS-IV Coding subtest, although examination of the interaction plot suggests the effect was primarily due to the performance of persons in the CCT with sham tDCS (Figure 4). The between group difference from CCT + sham was not significant at immediate follow-up (t [45.6] = 1.02, p = 0.57; d = 0.37) but approached significance at one-month follow-up (t [45.6] = 2.21, p = 0.08; d = 0.79).

**Figure 4.**
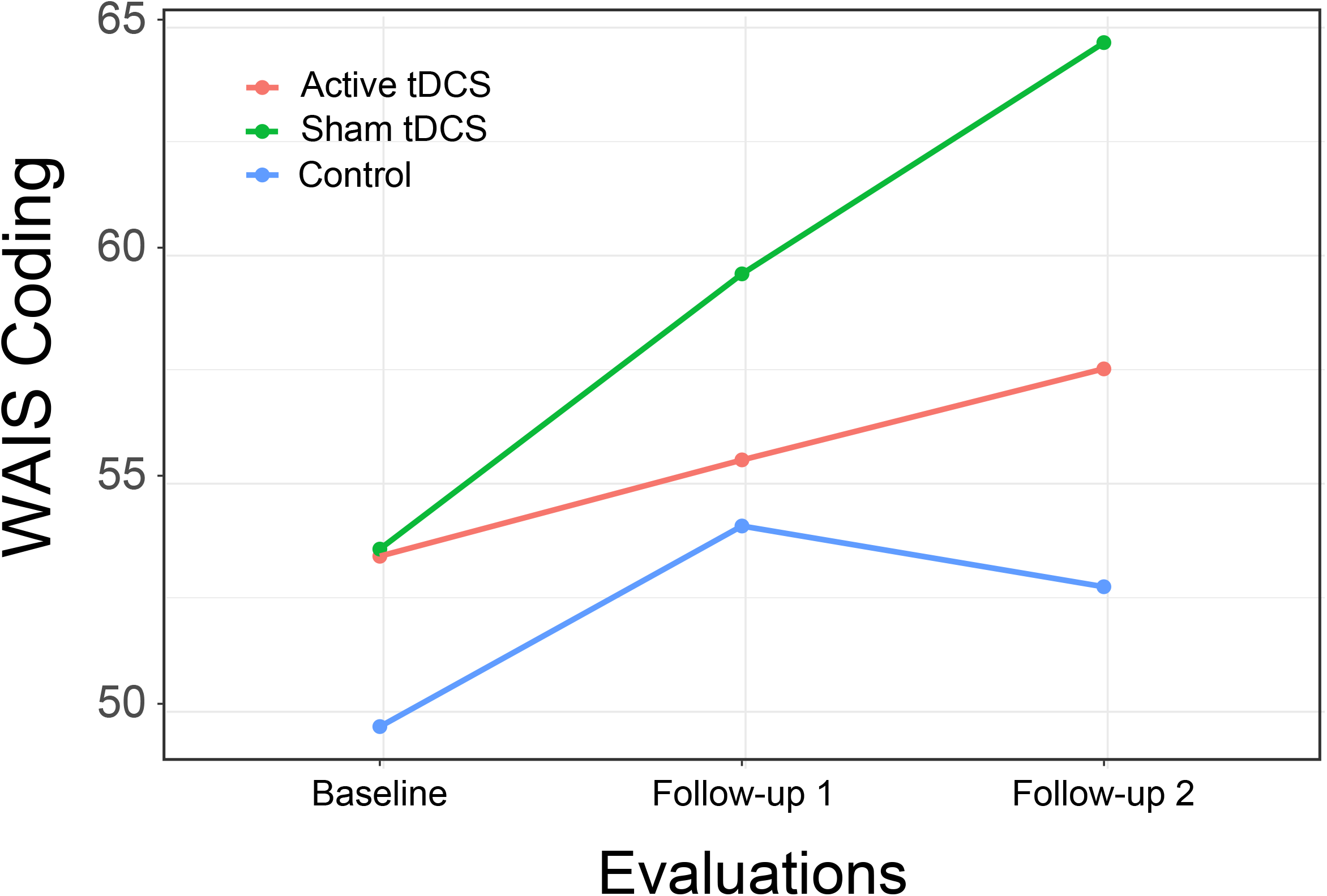
WAIS-IV Coding subtest scores by Group and Time

This pattern was again found in results for the HVLT total score, with a significant interaction of group by time resulting from differential improvement in both treatment groups, with significant difference between the CCT + active tDCS group and control at immediate follow-up (t [69.6] = 2.51, p = 0.04; d = 0.90) and a substantial but no longer significant difference at one-month follow-up (t [69.6] = 2.05, p = 0.11; d = 0.73; Figure 5).

**Figure 5.**
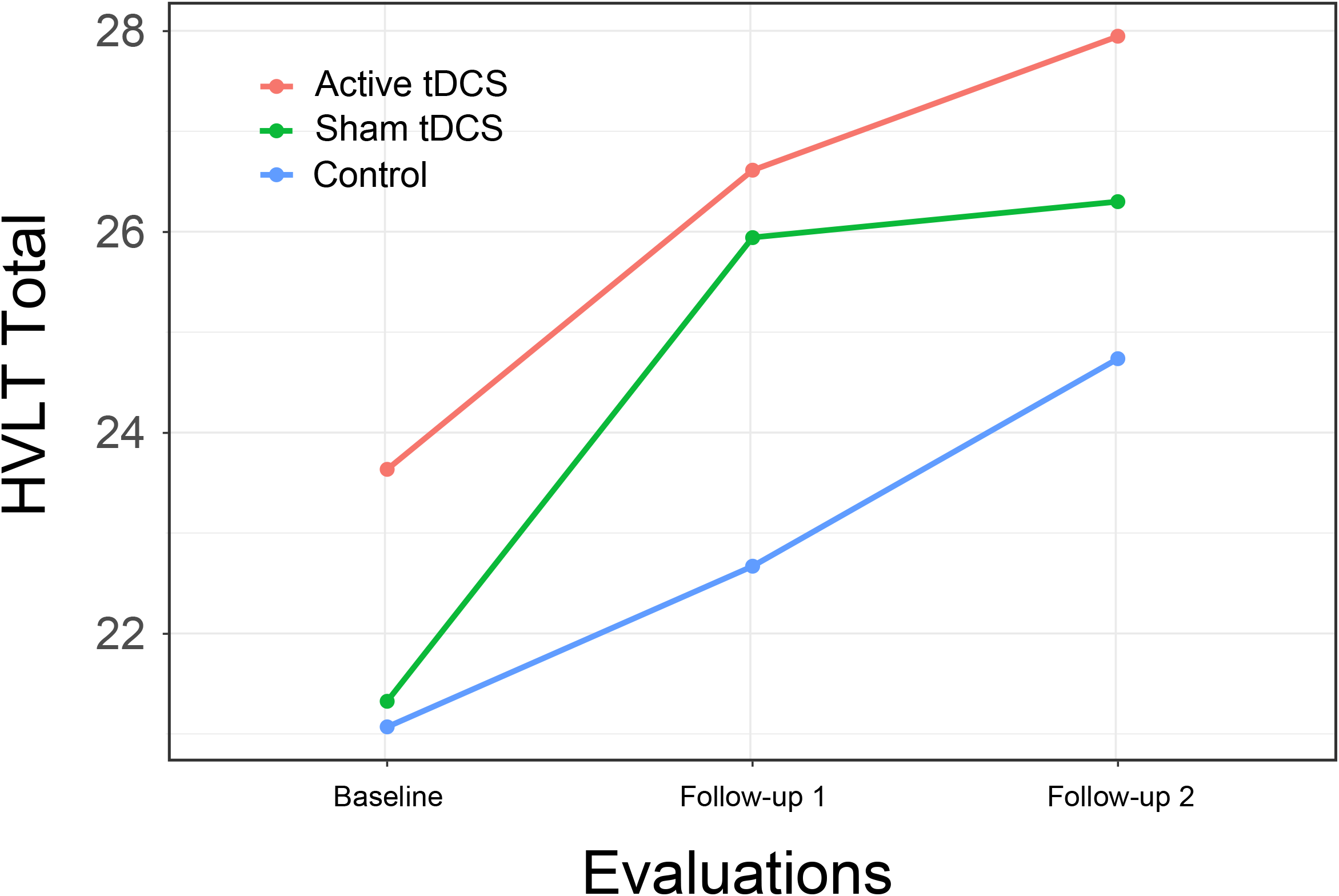
Hopkins Verbal Learning Test total score, Part B by Group and Time

Although we did not pose specific hypotheses for the effects of CCT with or without tDCS on executive function for this study, in exploratory analyses we evaluated the intervention’s effects on this domain with several measures. A significant interaction of group by time was found for the Stroop with a large effect size, but as with several other measures, the finding was related to substantial improvement in the CCT with sham stimulation group. Other interactions were again positive but nonsignificant, with effect sizes in the moderate range.

As with executive function, we did not propose specific hypotheses for the two functional measures included in the assessments, but explored the intervention’s effect on them. Results suggest a moderate but nonsignificant effect size for the MMT with a negligible effect size for the UPSA.

Given the large number of outcomes and our use of effect sizes to evaluate treatment effects, in an unplanned post-hoc analysis, we evaluated the overall impact of the study intervention based on the number of effect sizes for cognitive variables that were in the hypothesized direction (showing an interaction of group by time favoring one of the active treatment groups). The average effect size was 0.52, with 17 effects equal to or greater than 0.16, a small effect size [78]. The probability of this outcome compared to chance (equal distribution of positive effects across all groups) was significantly different (p = 0.04). It is thus improbable that the observed effects of the intervention were only due to chance. It should be acknowledged, that this analysis included positive effects for either treatment group and did not support the original hypothesis that the effects of CCT with active tDCS would be superior to CCT with sham tDCS.

#### 3.2.1 Self-report outcomes

Evaluations of treatment by time interactions of participant self-report on the PAOF subscales (PAOF Memory, χ2 [4] = 1.86, p = 0.76, d = 0.41; Cognition subscale, χ2 [4] = 0.37, p = 0.98, d = 0.18) and the CESD (χ2 [4] = 3.38, p = 0.50, d = 0.56) did not result suggest substantial between-group differences in response to the intervention. Examination of the interaction plots for the PAOF subscales (not presented) showed that mean scores for all three groups improved to a similar extent over the three evaluations. Participants reported similar levels of depression (CESD) across groups and evaluations, except participants in the active treatment group reported better mood at the one-month follow-up evaluation. Although the test of between groups differences were not significant, the within-group change from the immediate follow-up to the one-month follow-up for the CCT with active tDCS group approached significance and represented a large effect size (t [78.2] = 2.17, p = 0.08, d = 0.78).

Evaluation of participant ratings of how well they could think and remember resulted in an interaction that again approached statistical significance (χ2 [10] = 17.47, p = 0.06, d = 1.59). Examination of the interaction plot (Figure 6) suggests that persons in the active treatment group reported better ability to think over the final three training sessions, although between-group differences were not statistically significant (t [106.8] = 1.57, p = 0.26, d = 0.56).

**Figure 6.**
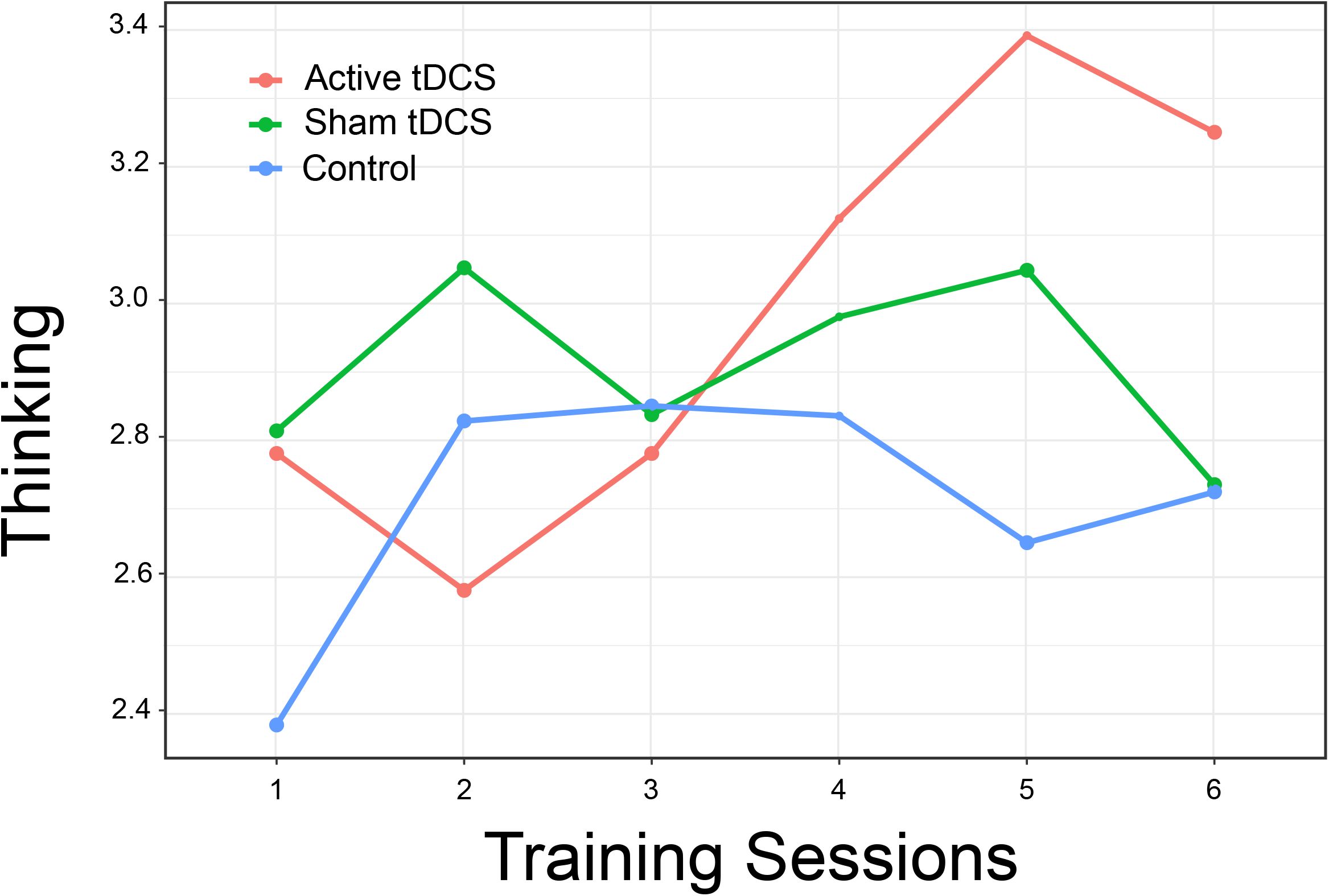
Participants’ Self-rated Thinking and Memory by Group and Training Session

## Discussion

This study’s goals were to assess the acceptability of CCT using a racing game to persons 50 years of age and older with HAND. The impact of adding tDCS to CCT was also evaluated. As we had a smaller than expected sample, we assessed outcomes not only through tests of statistical significance but also through calculation of effect sizes. Results showed that the majority of participants had positive views of the intervention and believed its benefits outweighed any risks. Ratings of its usefulness and usability were positive.

Our results clearly show that participants found the intervention acceptable. The majority of ratings on factors such as usefulness and ease of use were positive, and nearly all participants indicated that they felt the benefits of the intervention outweighed any drawbacks. Further exploration of ways to make the intervention even more positive for persons with HAND appear warranted.

Assessment of mental abilities before and after training suggested that the intervention had a positive effect on learning, memory, and motor speed compared to control. Although only a few outcomes were statistically significant or approached it, those which were significant were associated with large effect sizes. Outcomes with moderate or smaller effect sizes may thus have been nonsignificant due to low power related to the sample size rather than a lack of effect. A post-hoc evaluation of the probability of arriving at the observed set of effect sizes by chance suggested that our findings were not due to chance. Further, effect sizes obtained in this study are similar to those reported by other researchers, including in a meta-analysis of studies with older adults [86].

Consistent with other research, the combination of cognitive training and anodal tDCS at the left dorsolateral prefrontal cortex was associated with improvement in attention as measured by digit span [37, 87, 88]. Contrary to our hypothesis, however, there was no evidence of a substantial impact of the intervention on reaction time. Others have reported that tDCS may have a facilitating effect on reaction time with anodal stimulation over primary motor cortex, but Molero-Chamizo et al. found that the effect was time dependent [89]. Others have also failed to find an effect of tDCS [90, 91] on reaction time. Our hypothesis of an effect on reaction time was based primarily on the nature of the training task (a fast-paced computer game), but neither statistical tests nor inspection of mean plots by groups (not presented) suggested an effect for either of the active treatment groups.

We found more substantial improvements in measures of psychomotor speed, including a significant interaction of group by time for the WAIS-IV Coding subtest. Although the overall interaction for Trails B was not significant, inspection of group mean plots was consistent with relatively greater improvement on this measure for the two active treatment groups, and post-hoc between groups tests showed a significant difference between the control and CCT + active tDCS group after treatment while none was found at baseline.

A significant effect was also found for the Stroop Color-Word Test, a measure often interpreted as assessing a person’s ability to inhibit well-learned responses [92]. Others have observed a positive effect of tDCS on the Stroop task [93-95], while Frings et al. found that cathodal stimulation of the left DLPFC disrupted Stroop performance [96], with the negative effect of cathodal vs anodal stimulation expected.

We found limited support for the impact of the intervention on functional measures, with a moderate effect size but nonsignificant results for the Medication Management Test but no evidence of an effect on the UPSA. This is similar results reported by Vance et al. [97] who found limited effects of cognitive training on various measures of everyday function.

Self-report measures of mood or subjective cognitive functioning did not differ between groups, with one exception. The failure to find group differences may have been related to self-reported improvements across all groups, including the control. The one exception were self-reports of how well the participant perceived their thinking and memory, for which the CCT + active tDCS group gave substantially more positive reports over the last several sessions of the intervention.

The finding of possible treatment effects later during the training period suggests that the intervention’s effects may have continued after training ended, with continuing improvements at one-month follow-up in psychomotor speed (Trail Making Test, Part B; Figure 3) and verbal learning (Hopkins Verbal Learning Test; Figure 5). We also found an improvement in depression self-report (Center for Epidemiological Studies – Depression) at one-month follow-up. This possibly delayed effect of tDCS on mood was also reported by Li [98]. Given the evidence for improvement during the second week of training, it is possible that a more intensive and longer intervention might have resulted in greater effects. This possibility should be explored in future studies.

Strengths of the study include the single-blind sham-controlled design, which was effective in our pilot study as it has been in other studies [99]. We collected all data using a staff member who did not know the participant’s intervention assignment or by way of a computer. These measures reduced the likelihood of bias in outcomes due to experimenter effects. The characteristics of participants (age, gender, education level, cognitive function) made them similar to other persons who might have HAND and be able to benefit from the intervention. Another strength is the clear characterization of study participants with respect to concurrent medication use, although this in turn limited our ability to recruit participants.

This study’s limitations include the smaller than planned sample size. In spite of intensive recruiting efforts in the local community, including newspaper and online advertising, multiple contacts with community organizations and local infectious disease practitioners, and contacts with participants in previous studies we were not able to recruit the planned number of participants for the study. As shown in Figure 1, we were able to contact a number of potential participants that might have been adequate for planned sample size for the study, but exclusion due to validity concerns related to psychotropic medication use and safety concerns related to history of bipolar disorder, a large number of potential participants were not eligible. This limitation in turn may have affected the ability to test the statistical significance of outcomes, although in several cases when we found large effect sizes, we also found statistically significant results. Generalizing our results based only on observed effect sizes is a limitation. Finally, it should be acknowledged that while we found a number of positive effects on cognitive measures in the two CCT groups, the original hypothesis that CCT with active tDCS would be superior to CCT with sham tDCS was not supported.

Another limitation is that this study did not employ a double-blind controlled design, raising the possibility of bias caused by the investigators.

As noted above, we did several things to reduce the possibility of effects related to the unblinded experimenter. These included positioning the investigator and the tDCS device out of sight of the participant during stimulation, providing neutral suggestions about what the participants might experience during stimulation, and collecting most self-report data via computer assisted self-interview with the investigators out of the room. All baseline and follow-up assessments were conducted by an assessor blind to the participant’s treatment assignment or by way of a computer without the presence of a researcher. Thus, while we took a number of steps to reduce possible bias in the research design, they did not eliminate it.

HIV-related mental ability problems have implications for the functional status and quality of life for older persons with HAND. Our results, though limited, demonstrate the possibility that CCT with or without tDCS may have a positive impact on cognitive function. We found evidence of a moderate though nonsignificant effect of the intervention on a test of medication taking, a critically important skill for persons with HIV infection.

Future research should focus on continuing to explore the potential efficacy of CCT and tDCS with this population. A more detailed exploration of factors such as intensity and duration of stimulation and length and frequency of training sessions as well as the optimal timing of follow-up assessments may yield more effective treatment protocols with greater impacts on participants’ functional status.

## Data Availability

Anonymized data available from the investigators on request.

## 5 Conflict of Interest

The authors declare that they have no known competing financial interests or personal relationships that could have appeared to influence the work reported in this paper.

## 6 Author Contributions

RLO designed the study, obtained external funding, carried out the interventions, and wrote the draft of the manuscript. JK wrote a portion of the manuscript and reviewed the complete manuscript.

## 7 Funding

This work was supported by a grant from the US National Institutes on Aging to Dr. Ownby (grant R21AG056256).

## Notes

### Competing Interest Statement

The authors have declared no competing interest.

### Clinical Trial

NCT03440840

### Author Declarations

This protocol was approved by the Institutional Review Board of Nova Southeastern University (protocol number 2017-410).

### Summary of Updates

Rewritten to reduce similarity to our previous paper on the same topic.

